# How does childhood maltreatment influence cardiovascular disease? A sequential causal mediation analysis

**DOI:** 10.1101/2020.07.21.20158642

**Authors:** Ana G Soares, Laura D Howe, Jon Heron, Gemma Hammerton, Janet Rich-Edwards, Maria C Magnus, Sarah L Halligan, Abigail Fraser

**Author notes:** Correspondence to: Dr Ana Goncalves Soares, Population Health Sciences, Bristol Medical School, University of Bristol, Oakfield House, Oakfield Grove, Bristol, United Kingdom, BS8 2BN.

## Abstract

**Background:** Childhood maltreatment has been consistently associated with cardiovascular disease (CVD). However, the mechanisms of this relationship are not yet fully understood. We explored the relative contribution of anxiety/depression, smoking, body mass index (BMI) and inflammation (C-reactive protein, CRP) to the association between childhood maltreatment and CVD in men and women aged 40-69 years in the United Kingdom (UK).

**Methods:** We used data from 40,596 men and 59,511 women from UK Biobank. To estimate the indirect effects of childhood maltreatment (physical, sexual and emotional abuse, and emotional and physical neglect) on incident CVD via each of the mediators, we applied a sequential mediation approach.

**Results:** All forms of maltreatment were associated with increased CVD risk (hazard ratios (HR) ranging from 1.09 to 1.27). Together anxiety/depression, smoking, BMI and CRP mediated 26%-90% of the association between childhood maltreatment and CVD, and the contribution of these mediators differed by type of maltreatment and sex. Anxiety/depression mediated the largest proportion of the association of sexual abuse, emotional abuse and emotional neglect with CVD (accounting for 16%-43% of the total effect), especially in women. In men, BMI contributed the most to the indirect effect of associations of physical abuse and physical neglect with CVD; in women anxiety/depression and BMI had similar contributions.

**Conclusions:** These findings add to the understanding of how childhood maltreatment affects CVD risk and identify modifiable mediating factors which could potentially reduce the burden of CVD in people exposed to maltreatment in early life.

## Introduction

There is a robust association between childhood maltreatment and cardiovascular disease (CVD) risk in later life ^(1)^, but the mechanisms underlying this relationship are not fully understood ^(2)^. Three main pathways are commonly postulated: behavioural (e.g. smoking, unhealthy diet/overeating and physical inactivity), mental health (e.g. mood, anxiety and depressive disorders) and biological factors (alterations in the immune, metabolic, neuroendocrine, and autonomic nervous systems) ^(2-4)^. However, few studies have explicitly tested and/or quantified these mechanisms with comprehensive mediation models ^(2, 5)^, and it is not clear to what extent the association between childhood maltreatment and CVD acts through these proposed pathways or via other mechanisms.

Most studies have assessed childhood adversities as a summary score ^(5, 6)^, and to the best of our knowledge, no study has explored whether the mechanisms of this association differ according to the type of maltreatment suffered. Moreover, there is some evidence of sex differences in the association between childhood maltreatment and CVD, though with inconsistent patterns ^(1, 7, 8)^. The few studies that explored potential sex-specific mechanisms found similar results in men and women ^(3, 7)^.

In this study, we used data from UK Biobank and applied sequential causal mediation analysis ^(9)^ using g-computation ^(10)^ to estimate the relative contribution of anxiety/depression, smoking, body mass index (BMI) and inflammation to the association between childhood maltreatment and CVD. This approach allows us to estimate the combined and individual contributions of each mediator, based on a causal order, whilst handling intermediate confounding. We also explored possible differences by sex and by type of maltreatment.

## Methods

UK Biobank recruited 502,524 participants aged 40-69 years between 2006 and 2010 ^(11)^. In 2016, participants who provided an email address (n=339,092) were invited to complete an online mental health questionnaire, which included questions about their experiences of maltreatment in childhood ^(12)^. In total, 158,835 participants responded the online questionnaire (47% of those emailed) ^(12)^, and 157,311 responded to at least one question about childhood maltreatment. We excluded participants with CVD at baseline, when information of mediators was collected (details in supplementary material). We also excluded those with incomplete data on mediators and confounders. 40,596 men and 59,511 women were included in the analyses (eFigure 1). The majority (91.3%) of the eligible sample had complete data on all mediators and confounders, and after accounting for covariates, missingness was not associated with the outcome or the exposures, apart from a lower likelihood of reporting physical neglect in those with complete data (eTable 1). In this scenario, complete case analysis is unlikely to introduce bias ^(13)^.

Childhood maltreatment was assessed using the Childhood Trauma Screener ^(14)^, and classified into: physical, sexual and emotional abuse, and emotional and physical neglect. CVD incident cases were extracted from hospital and death registers and coded according to the International Classification of Diseases (ICD-10 codes: I00-I99). We considered anxiety/depression, smoking, BMI and inflammation, indexed by C-reactive protein (CRP), all assessed at baseline, as potential mediators.

Information on age, year of birth, ethnicity, maternal smoking around birth, number of siblings, and family history of CVD was obtained at recruitment and used as baseline confounders in our analysis. As UK Biobank lacks information on childhood socioeconomic position (SEP), we used maternal smoking, number of siblings, and family history of CVD as proxies, considering their socioeconomic patterns ^(15)^.

Given the long lag time between exposure to childhood maltreatment and the emergence and/or assessment of the potential mediators, other factors, such as adult socioeconomic position ^(16)^, might be affected by childhood maltreatment but also confound the association between potential mediators and CVD (i.e. intermediate confounding). We considered Townsend deprivation index and educational attainment as intermediate confounders.

Further details on how the exposures, mediators and confounders were assessed and categorised are presented in supplementary material.

### Statistical analysis

All analyses were conducted using Stata version 15.1 (Statacorp, College Station, TX). We used Cox proportional hazards models to estimate hazard ratios (HR) and 95% confidence intervals (95% CIs) for the associations between childhood maltreatment and CVD and between the mediators and CVD. Contributions to risk were censored at the first CVD event, death for any reason or end of follow-up, on 31 March 2017. The time scale for the Cox model was calendar year. We used linear regression to assess the associations of childhood maltreatment with both BMI and CRP, and logistic regression to assess the associations with anxiety/depression and smoking. Analyses were performed unadjusted and adjusted for potential confounders; baseline confounders were used for exposure-outcome and exposureediator associations, and baseline and intermediate confounders were used for mediator outcome associations.

### Mediation analysis

The proposed causal diagram for the relationships between childhood maltreatment and CVD is shown in Figure 1. We assumed that childhood maltreatment leads to anxiety/depression ^(17)^, which influences smoking ^(18)^, then adiposity ^(19)^, and results in changes in inflammatory markers ^(20, 21)^. Adult SEP, which is affected by childhood maltreatment ^(16)^, might also confound the association between childhood maltreatment and behavioural, mental health and biological factors ^(15)^, and therefore was considered an intermediate confounder.

**Figure 1.**
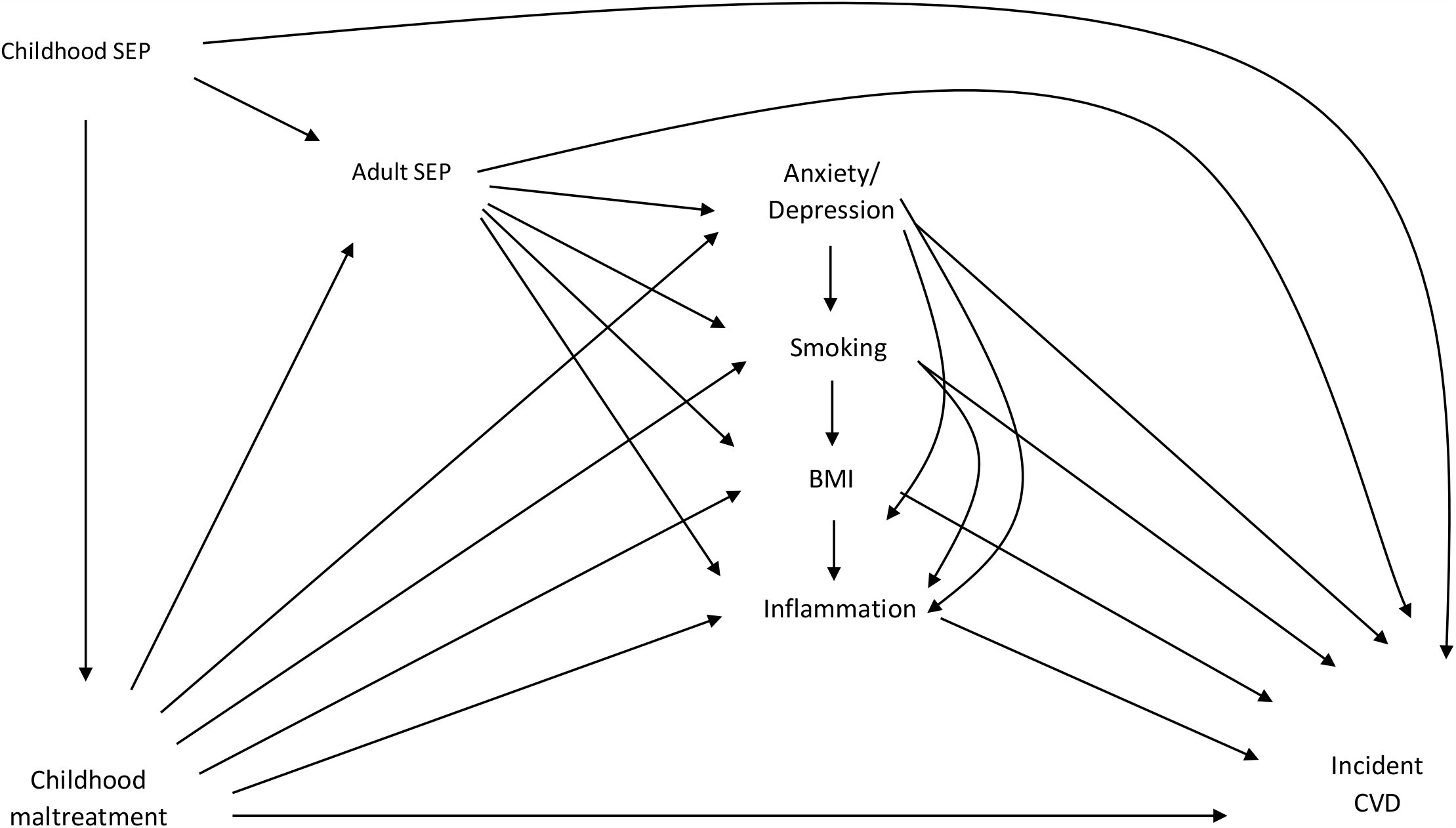
Assumed causal diagram of the association between childhood maltreatment and cardiovascular disease (CVD) BMI: body mass index; CVD: cardiovascular disease; SEP: socioeconomic position

To estimate direct and indirect effects, the following assumptions regarding confounding are needed: a) no unmeasured confounding for the exposure-outcome relationship, b) no unmeasured confounding for the mediator-outcome relationship, c) no unmeasured confounding for the exposure-mediator relationship, and d) no effects of the exposure that confounds the mediator-outcome relationship ^(9)^. To decompose the effect of each type of childhood maltreatment on CVD risk into natural indirect effect (NIE, i.e. acting through the mediators) and natural direct effect (NDE, i.e. not acting through the mediators), we used parametric g-computation using Monte Carlo simulations, performed using the Stata command *gformula* ^(10)^. This approach, which uses counterfactual definitions of direct and indirect effects, can handle intermediate confounding, and therefore there is no need to make assumption d, which is needed when using standard methods for mediation ^(10)^. We considered the natural direct effect (NDE) and natural indirect effect (NIE) as decompositions of the total causal effect (TCE); *gformula* implements logistic regression for time-to-event outcomes and therefore estimates correspond to log odds ratios (ORs). We exponentiated the results and present ORs.

As we were interested in the combined contribution of the set of mediators, but also of each individual mediator, we performed sequential mediation analysis ^(9, 22)^. This approach allows for mediation analysis using multiple related mediators, based on the assumption of a causal ordering of the mediators. We estimated four models (Figure 2). In Model 1, we estimated the NIE through anxiety/depression; this includes pathways that act through anxiety/depression and any of its effects (descendants) but does not include those that act solely through smoking, BMI and/or CRP (or other pathways). In Model 2, the NIE through both anxiety/depression and smoking was estimated, which includes their causal descendants, but does not include any other pathways, including paths that act only via BMI and/or CRP; the difference between the proportions mediated in Model 2 and Model 1 corresponds to the proportion mediated by smoking beyond anxiety/depression alone. In Model 3, the NIE through anxiety/depression, smoking and BMI was estimated, which includes their causal descendants, but does not include the paths that act solely through CRP; the difference between the proportions mediated in Model 3 and Model 2 corresponds to the mediating effect of BMI beyond the effect of both anxiety/depression and smoking. Finally, in Model 4, the NIE of all potential mediators was estimated, and the difference between the proportion mediated in Model 4 and the proportion mediated in Model 3 corresponds to the mediating effect of CRP beyond anxiety/depression, smoking and BMI. Exposure-mediator interactions were included sequentially (i.e. interaction between maltreatment and anxiety/depression was included in Model 1, interactions between maltreatment and anxiety/depression and between maltreatment and smoking were included in Model 2, and so on). We did not include mediator-mediator interactions, as these would hinder comparability across models ^(9)^. Bootstrapping with 1,000 replications was used to calculate 95% CIs. The number of Monte Carlo simulations was equivalent to the sample size of each model. The proportion mediated was calculated as 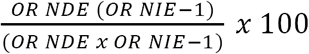.

**Figure 2.**
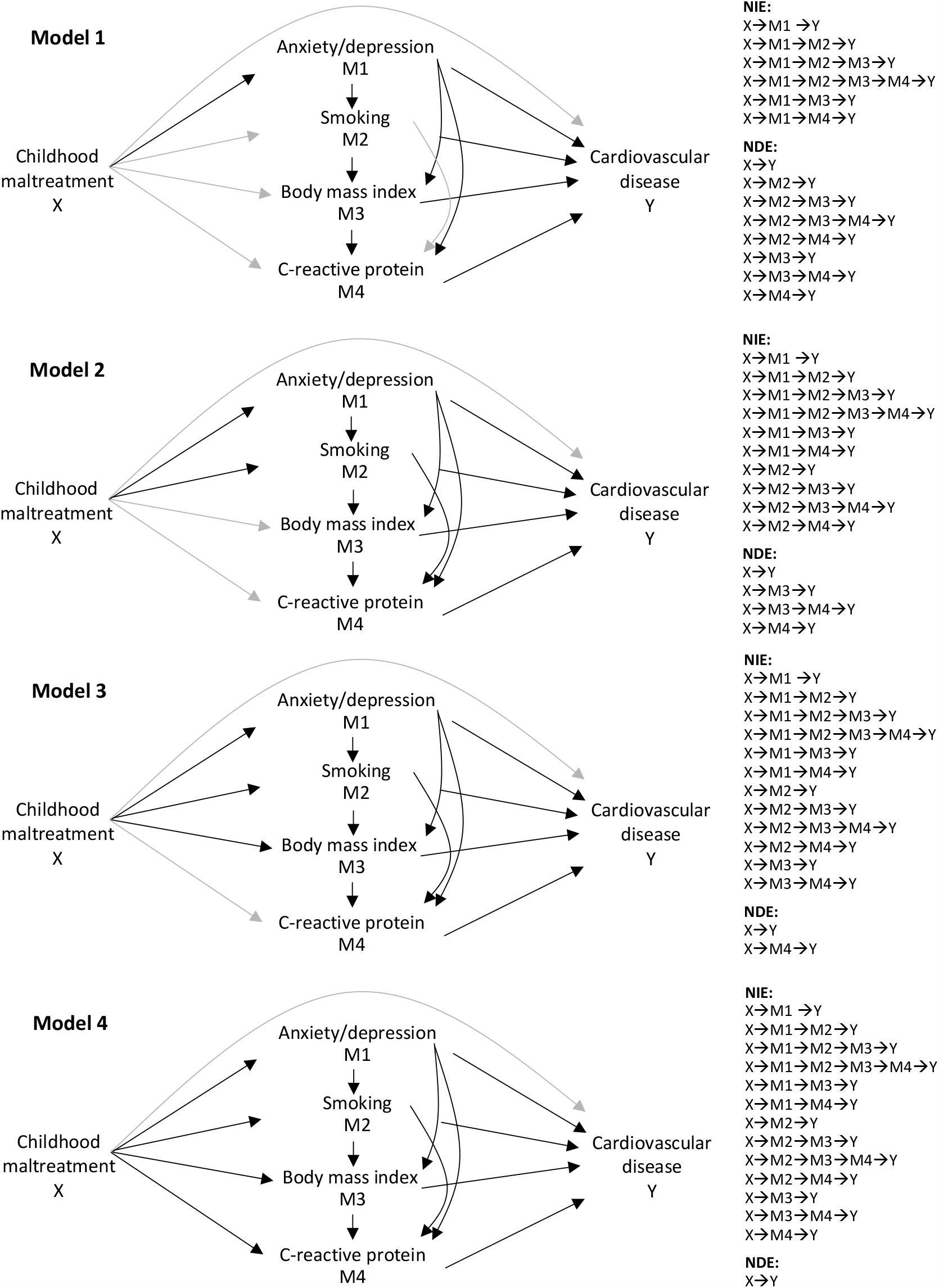
Simplified causal diagrams illustrating estimated paths in Models 1-4; the natural direct effect (NDE) is illustrated by the grey lines and the natural indirect effect (NIE) by the black lines.

All analyses were performed stratified by sex, given previously reported sex differences in the association between childhood maltreatment and CVD and the scarcity of studies exploring sex differences in the mechanisms of this association. We compared the NDE and NIE between men and women by examining the point estimates and their 95% confidence intervals.

### Sensitivity analysis

Sequential mediation analysis requires a causal ordering of the mediators ^(9)^. However, the temporal and causal order is arguable for this set of mediators. Observational studies suggest a bidirectional relationship between depression and BMI ^(23)^, and Mendelian randomization studies (i.e. using genetic variants as instrumental variables) support a causal role for higher BMI on the likelihood of depression, which may also be bidirectional ^(24, 25)^. There is evidence of a causal effect of smoking on BMI ^(26, 27)^, but also of BMI on smoking ^(28, 29)^. Furthermore, smoking is a risk factor for depression, and the genetic liability for depression increases lifetime smoking ^(30)^.

As the order in which mediators are included in the statistical models could potentially influence results, we investigated in sensitivity analysis the robustness of findings to the order of inclusion. We first estimated the NIE through BMI, then added anxiety/depression, then smoking, and finally CRP. The conceptualized causal diagram for these relationships is shown in eFigure 2, and the simplified causal diagram illustrating the estimated paths in each model is presented in eFigure 3.

## Results

Characteristics of participants without CVD at baseline who were included and excluded (due to missing data on any of exposures/confounders/mediators) from analyses are shown in eTable 2. Included participants were more likely to be female and White/British. Men who were included in the analyses were slightly older, whilst women were younger. The participants included were also more likely to experience less deprivation, to have higher education, fewer siblings, lower prevalence of maternal smoking, current smoking and anxiety/depression, and higher frequency of alcohol intake than those not included in the analyses. Additionally, participants included were more likely to have lower BMI, CRP, systolic and diastolic blood pressures, use of blood pressure medication, and lower rates of CVD than those not included.

Both physical abuse and emotional neglect were the most prevalent types of childhood maltreatment in men (20.8%), and emotional neglect was the most prevalent in women (22.3%) (eFigure 4). The prevalence of all types of maltreatment was higher in women, except for physical abuse, which was higher in men. The incidence rate of CVD was 20.9 per 1,000 person-years (95% CI 20.4; 21.4) in men and 14.0 per 1,000 person-years (95% CI 13.7; 14.3) in women.

After adjusting for baseline confounders, all types of childhood maltreatment were associated with higher risk of incident CVD in both men and women, with HR varying from 1.09 to 1.27 (eTable 3). Associations were overall similar for all types of maltreatment but were slightly stronger for emotional abuse, especially in women. There was little evidence of sex differences, except for a stronger association between emotional neglect and CVD in women.

Childhood maltreatment was associated with higher BMI and CRP and with higher odds of smoking and anxiety/depression in both sexes (eTable 4). The exceptions were for the associations between emotional neglect and BMI and between physical neglect and smoking in men. There was some evidence of stronger associations in women between childhood maltreatment and potential mediators. All mediators were associated with higher risk of incident CVD (eTable 5). The associations did not differ by sex, except for the association between anxiety/depression and CVD, which was stronger in women.

Tables 1 and 2 show the TCE, NDE and NIE of the associations between childhood maltreatment and CVD in men and women, respectively, as well as the proportion of the total effect explained by the mediators individually and in combination. Anxiety/depression, smoking, BMI and CRP mediated 26% to 90% of the associations between childhood maltreatment and CVD. Overall, these mediators explained a greater proportion of the associations in women; for example, the four mediators together explained 90% of the association between sexual abuse and CVD in women, compared to 34% in men.

**Table 1.**
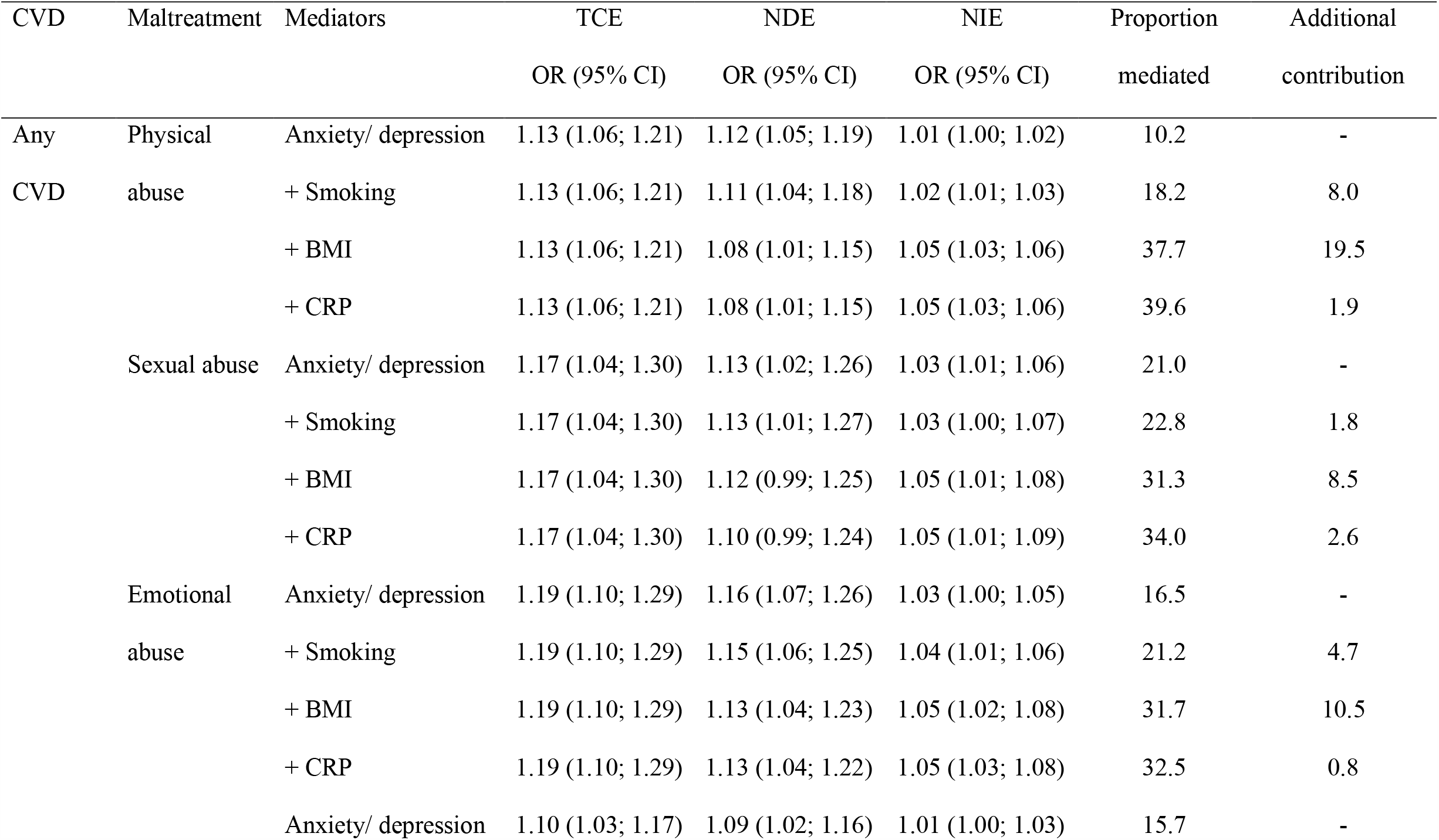
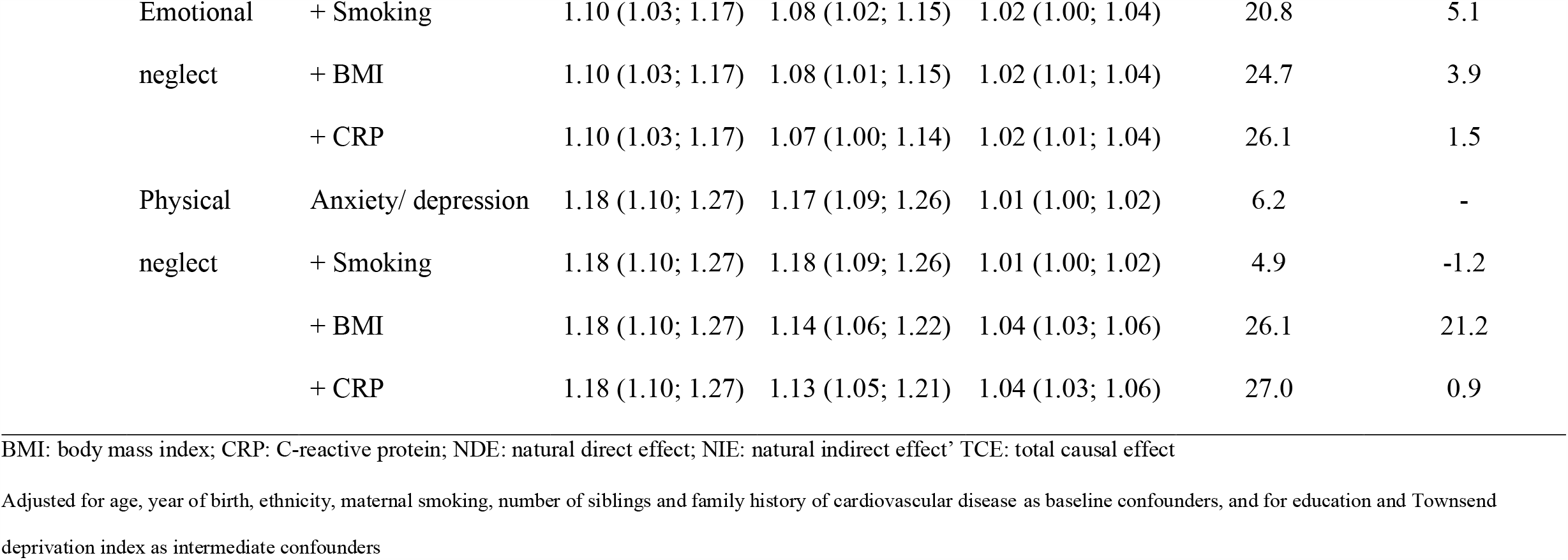
Estimated odds ratios for the association between childhood maltreatment and cardiovascular disease in men.

| CVD | Maltreatment | Mediators | TCE | NDE | NIE | Proportion | Additional |
| --- | --- | --- | --- | --- | --- | --- | --- |
|  |  |  | OR (95% CI) | OR (95% CI) | OR (95% CI) | mediated | contribution |
| Any | Physical | Anxiety/ depression | 1.13 (1.06; 1.21) | 1.12 (1.05; 1.19) | 1.01 (1.00; 1.02) | 10.2 | - |
| CVD | abuse | + Smoking | 1.13 (1.06; 1.21) | 1.11 (1.04; 1.18) | 1.02 (1.01; 1.03) | 18.2 | 8.0 |
|  |  | + BMI | 1.13 (1.06; 1.21) | 1.08 (1.01; 1.15) | 1.05 (1.03; 1.06) | 37.7 | 19.5 |
|  |  | + CRP | 1.13 (1.06; 1.21) | 1.08 (1.01; 1.15) | 1.05 (1.03; 1.06) | 39.6 | 1.9 |
|  | Sexual abuse | Anxiety/ depression | 1.17 (1.04; 1.30) | 1.13 (1.02; 1.26) | 1.03 (1.01; 1.06) | 21.0 | - |
|  |  | + Smoking | 1.17 (1.04; 1.30) | 1.13 (1.01; 1.27) | 1.03 (1.00; 1.07) | 22.8 | 1.8 |
|  |  | + BMI | 1.17 (1.04; 1.30) | 1.12 (0.99; 1.25) | 1.05 (1.01; 1.08) | 31.3 | 8.5 |
|  |  | + CRP | 1.17 (1.04; 1.30) | 1.10 (0.99; 1.24) | 1.05 (1.01; 1.09) | 34.0 | 2.6 |
|  | Emotional abuse | Anxiety/ depression | 1.19 (1.10; 1.29) | 1.16 (1.07; 1.26) | 1.03 (1.00; 1.05) | 16.5 | - |
|  |  | + Smoking | 1.19 (1.10; 1.29) | 1.15 (1.06; 1.25) | 1.04 (1.01; 1.06) | 21.2 | 4.7 |
|  |  | + BMI | 1.19 (1.10; 1.29) | 1.13 (1.04; 1.23) | 1.05 (1.02; 1.08) | 31.7 | 10.5 |
|  |  | + CRP | 1.19 (1.10; 1.29) | 1.13 (1.04; 1.22) | 1.05 (1.03; 1.08) | 32.5 | 0.8 |
|  |  | Anxiety/ depression | 1.10 (1.03; 1.17) | 1.09 (1.02; 1.16) | 1.01 (1.00; 1.03) | 15.7 | - |

|  |  |  |  |  |  |  |
| --- | --- | --- | --- | --- | --- | --- |
| Emotional | + Smoking | 1.10 (1.03; 1.17) | 1.08 (1.02; 1.15) | 1.02 (1.00; 1.04) | 20.8 | 5.1 |
| neglect | + BMI | 1.10 (1.03; 1.17) | 1.08 (1.01; 1.15) | 1.02 (1.01; 1.04) | 24.7 | 3.9 |
|  | + CRP | 1.10 (1.03; 1.17) | 1.07 (1.00; 1.14) | 1.02 (1.01; 1.04) | 26.1 | 1.5 |
| Physical | Anxiety/ depression | 1.18 (1.10; 1.27) | 1.17 (1.09; 1.26) | 1.01 (1.00; 1.02) | 6.2 | - |
| neglect | + Smoking | 1.18 (1.10; 1.27) | 1.18 (1.09; 1.26) | 1.01 (1.00; 1.02) | 4.9 | -1.2 |
|  | + BMI | 1.18 (1.10; 1.27) | 1.14 (1.06; 1.22) | 1.04 (1.03; 1.06) | 26.1 | 21.2 |
|  | + CRP | 1.18 (1.10; 1.27) | 1.13 (1.05; 1.21) | 1.04 (1.03; 1.06) | 27.0 | 0.9 |

BMI: body mass index; CRP: C-reactive protein; NDE: natural direct effect; NIE: natural indirect effect' TCE: total causal effect
Adjusted for age, year of birth, ethnicity, maternal smoking, number of siblings and family history of cardiovascular disease as baseline confounders, and for education and Townsend deprivation index as intermediate confounders

**Table 2.**
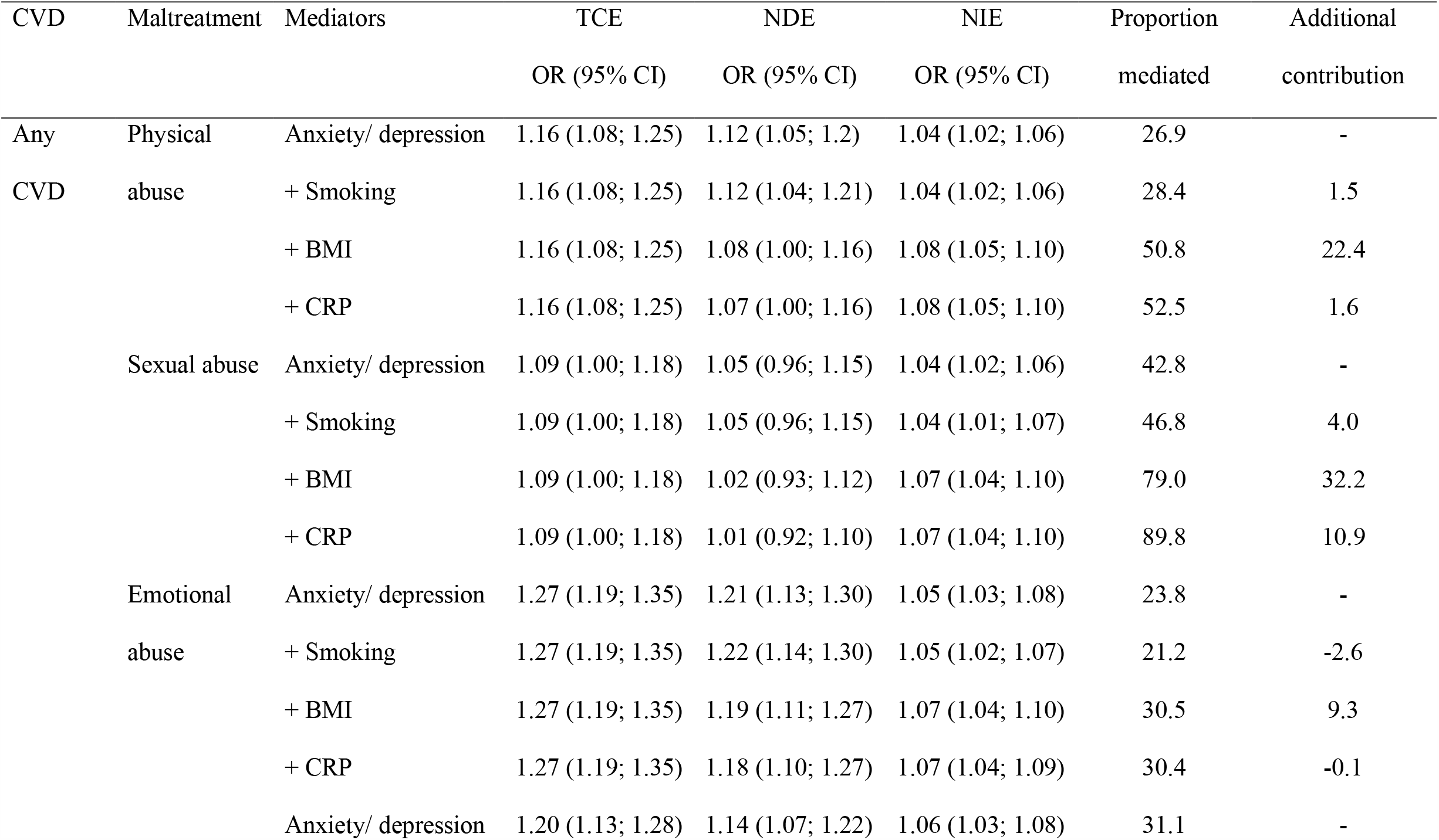
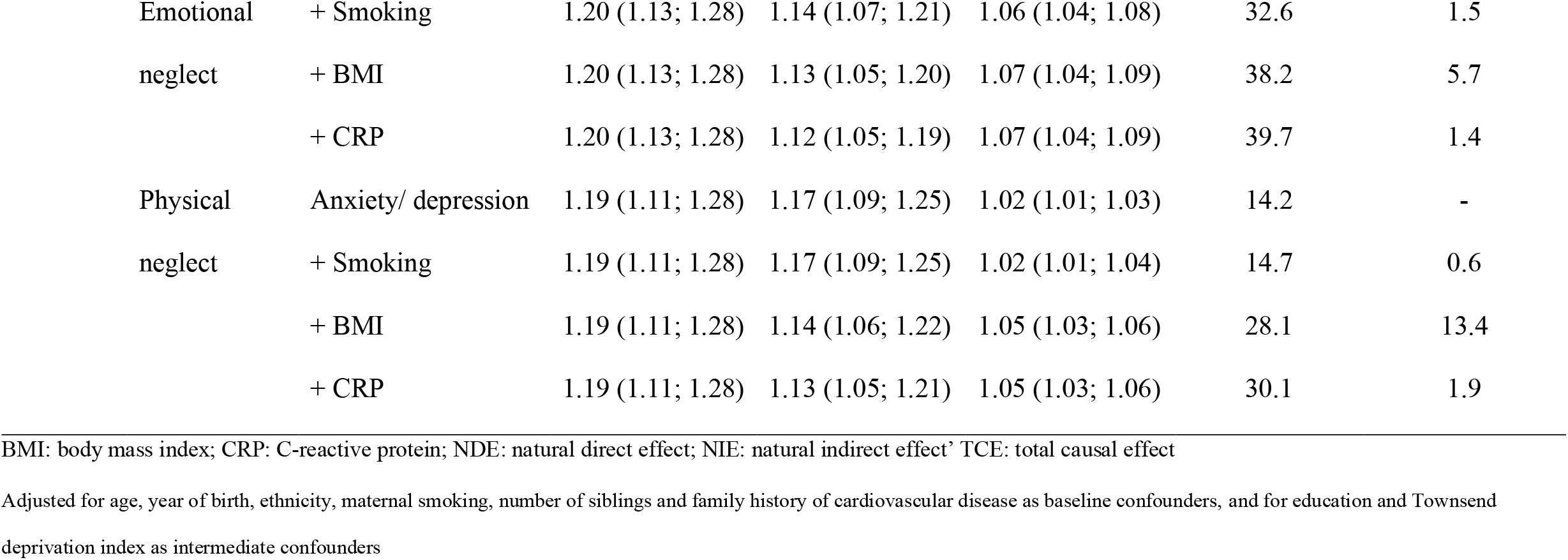
Estimated odds ratios for the association between childhood maltreatment and cardiovascular disease in women.

| CVD | Maltreatment | Mediators | TCE | NDE | NIE | Proportion | Additional |
| --- | --- | --- | --- | --- | --- | --- | --- |
|  |  |  | OR (95% CI) | OR (95% CI) | OR (95% CI) | mediated | contribution |
| Any | Physical | Anxiety/ depression | 1.16 (1.08; 1.25) | 1.12 (1.05; 1.2) | 1.04 (1.02; 1.06) | 26.9 | - |
| CVD | abuse | + Smoking | 1.16 (1.08; 1.25) | 1.12 (1.04; 1.21) | 1.04 (1.02; 1.06) | 28.4 | 1.5 |
|  |  | + BMI | 1.16 (1.08; 1.25) | 1.08 (1.00; 1.16) | 1.08 (1.05; 1.10) | 50.8 | 22.4 |
|  |  | + CRP | 1.16 (1.08; 1.25) | 1.07 (1.00; 1.16) | 1.08 (1.05; 1.10) | 52.5 | 1.6 |
|  | Sexual abuse | Anxiety/ depression | 1.09 (1.00; 1.18) | 1.05 (0.96; 1.15) | 1.04 (1.02; 1.06) | 42.8 | - |
|  |  | + Smoking | 1.09 (1.00; 1.18) | 1.05 (0.96; 1.15) | 1.04 (1.01; 1.07) | 46.8 | 4.0 |
|  |  | + BMI | 1.09 (1.00; 1.18) | 1.02 (0.93; 1.12) | 1.07 (1.04; 1.10) | 79.0 | 32.2 |
|  |  | + CRP | 1.09 (1.00; 1.18) | 1.01 (0.92; 1.10) | 1.07 (1.04; 1.10) | 89.8 | 10.9 |
|  | Emotional<br>abuse | Anxiety/ depression | 1.27 (1.19; 1.35) | 1.21 (1.13; 1.30) | 1.05 (1.03; 1.08) | 23.8 | - |
|  |  | + Smoking | 1.27 (1.19; 1.35) | 1.22 (1.14; 1.30) | 1.05 (1.02; 1.07) | 21.2 | -2.6 |
|  |  | + BMI | 1.27 (1.19; 1.35) | 1.19 (1.11; 1.27) | 1.07 (1.04; 1.10) | 30.5 | 9.3 |
|  |  | + CRP | 1.27 (1.19; 1.35) | 1.18 (1.10; 1.27) | 1.07 (1.04; 1.09) | 30.4 | -0.1 |
|  |  | Anxiety/ depression | 1.20 (1.13; 1.28) | 1.14 (1.07; 1.22) | 1.06 (1.03; 1.08) | 31.1 | - |

|  |  |  |  |  |  |  |
| --- | --- | --- | --- | --- | --- | --- |
| Emotional | + Smoking | 1.20 (1.13; 1.28) | 1.14 (1.07; 1.21) | 1.06 (1.04; 1.08) | 32.6 | 1.5 |
| neglect | + BMI | 1.20 (1.13; 1.28) | 1.13 (1.05; 1.20) | 1.07 (1.04; 1.09) | 38.2 | 5.7 |
|  | + CRP | 1.20 (1.13; 1.28) | 1.12 (1.05; 1.19) | 1.07 (1.04; 1.09) | 39.7 | 1.4 |
| Physical | Anxiety/ depression | 1.19 (1.11; 1.28) | 1.17 (1.09; 1.25) | 1.02 (1.01; 1.03) | 14.2 | - |
| neglect | + Smoking | 1.19 (1.11; 1.28) | 1.17 (1.09; 1.25) | 1.02 (1.01; 1.04) | 14.7 | 0.6 |
|  | + BMI | 1.19 (1.11; 1.28) | 1.14 (1.06; 1.22) | 1.05 (1.03; 1.06) | 28.1 | 13.4 |
|  | + CRP | 1.19 (1.11; 1.28) | 1.13 (1.05; 1.21) | 1.05 (1.03; 1.06) | 30.1 | 1.9 |

BMI: body mass index; CRP: C-reactive protein; NDE: natural direct effect; NIE: natural indirect effect' TCE: total causal effect
Adjusted for age, year of birth, ethnicity, maternal smoking, number of siblings and family history of cardiovascular disease as baseline confounders, and for education and Townsend deprivation index as intermediate confounders

The contribution of the mediators to the association between childhood maltreatment and CVD varied by type of maltreatment and sex. For sexual abuse, emotional abuse and emotional neglect, anxiety/depression explained the highest proportion of the indirect effect, accounting for 16%-43% of the total effect, with higher proportions observed in women (Tables 1 and 2, Figure 3). For physical abuse and physical neglect, BMI contributed the most to the indirect effect in men, and in women both anxiety/depression and BMI had similar contributions.

**Figure 3.**
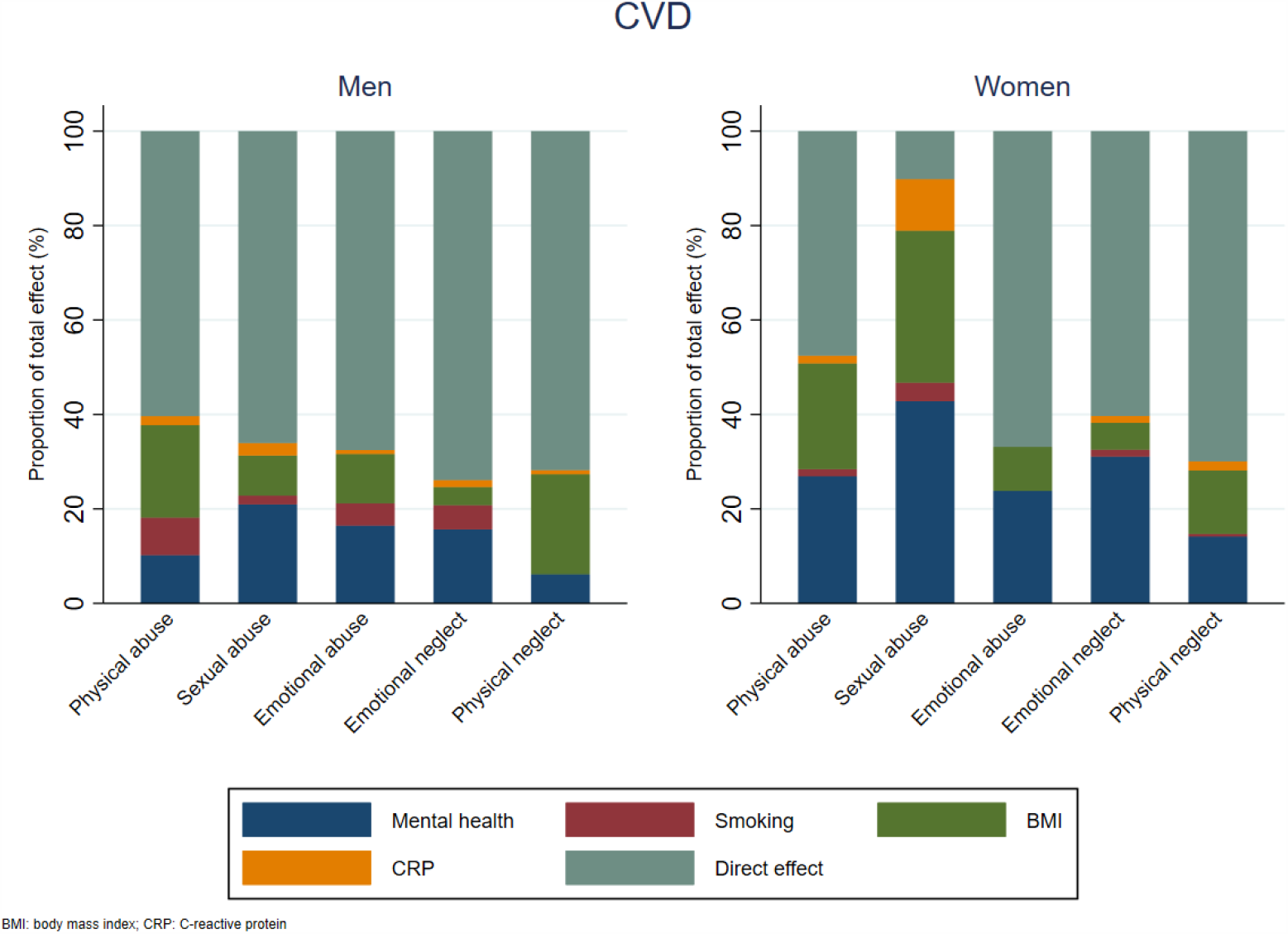
Proportion mediated by mental health, smoking, body mass index and inflammation in men and women in the association between childhood maltreatment and cardiovascular disease.

### Sensitivity analysis

When we first included BMI, then anxiety/depression, then smoking, and finally CRP, the proportion explained by each mediator changed only slightly (eTables 6 and 7, eFigure 5) and the conclusions regarding the main mediators of the associations between childhood maltreatment and CVD did not change.

## Discussion

This study assessed the role of anxiety/depression, smoking, BMI and inflammation in the relationship between childhood maltreatment and incident CVD in men and women aged 40-69 years in the UK using a sequential mediation approach. We showed that together these factors mediated 26%-90% of the associations between childhood maltreatment and CVD. A higher proportion of the association between childhood maltreatment and CVD was explained by this set of mediators in women than in men, and anxiety/depression seemed to have a particularly important role for women.

Longitudinal studies have shown that mental health has an important mediating effect in the association between childhood adversity and CVD alongside traditional CVD risk factors, such as smoking, unhealthy diet, obesity and physical inactivity ^(3, 5, 7, 31)^. Similarly, we found that mental health explained more of the association of sexual abuse, emotional abuse and emotional neglect with CVD than BMI, smoking and inflammation, particularly in women. Most previous studies assessed the mediating role of mental health independently of other health behaviours, without taking the relationships between them into account, including a recent study using UK Biobank that showed that depressive symptoms mediated 56% of the association between a summary score of childhood maltreatment and CVD in men and women combined ^(5)^. A USA longitudinal study of young adults (mean age 30y) used path analysis to consider depressive symptoms along with other factors, and found that depressive symptoms did not mediate the effect of childhood adversities on CVD risk factors ^(32)^. In our study, the importance of anxiety/depression as a mediator of the association between childhood maltreatment and CVD was consistent even when the order of the mediators was changed.

Women have a greater burden of childhood maltreatment ^(33)^, as well as anxiety and depression ^(34)^, and depression has been linked to CVD in both men and women ^(35, 36)^, including in prospective studies ^(37, 38)^. The association between depression and CVD has been shown to be stronger in women in some ^(39)^ but not all studies ^(40)^. Several mechanisms may link anxiety/depression to CVD; alterations in the autonomic nervous system, platelet receptors and function, coagulopathic factors, proinflammatory cytokines, endothelial function, neurohormonal factors, genetic linkages, and adverse health behaviours ^(35)^. However, causal evidence is still limited. A recent study using Mendelian randomization suggests that depression and CVD comorbidity arises largely from shared environmental factors, and that triglycerides and inflammation are likely to be causally related to depression ^(41)^.

BMI was also an important mediator in the relationship between childhood maltreatment and CVD. It seemed to be a major explanatory factor for the relationship of physical abuse and neglect with CVD in men, and its contribution to the association of physical abuse and neglect with CVD in women was comparable to that of anxiety/depression. Childhood maltreatment has been robustly associated with higher BMI and risk of obesity in adults ^(42, 43)^, and there is strong evidence of a causal association between higher BMI and CVD ^(44)^. Sex differences in the association between BMI and CVD are not consistent ^(42, 45, 46)^, and we found that the contribution of BMI to the association between childhood maltreatment and CVD differed in men and women.

Evidence suggests that childhood maltreatment is associated with inflammation ^(47-49)^, and that inflammation mediates the association between childhood abuse and hypertension ^(50)^. In women, inflammation explained more of the indirect effect than smoking. Although CRP is not causally related to CVD ^(51)^, its production is stimulated by IL-6, which likely has a causal effect on CVD ^(52)^. CRP is affected by both BMI ^(48, 53)^ and smoking ^(20)^, and these might have already captured most of the effect of CRP, which was entered last into the models. This might also explain the relatively small additional contribution of smoking; its indirect effect might have already been captured by anxiety/depression ^(18)^, especially in women ^(54)^. Smoking and BMI have bidirectional effects ^(28, 29)^, but they might also reflect two separate coping mechanisms, and individual differences might influence whether individuals are more likely to adopt one over the other. Eating disorders, for example, are more common in women, and differences by type of maltreatment and sex have been shown in the association between childhood maltreatment and eating disorders ^(55)^. Smoking, on the other hand, is more prevalent in men ^(56)^, and sex differences in the association between childhood maltreatment and smoking have been observed ^(57)^.

### Strengths and limitations

As most large longitudinal studies, we used self-reported retrospective measures of childhood maltreatment, which precludes robust measurements of severity and timing of the exposures reported and might be affected by recall bias and/or measurement error ^(58, 59)^. Although there is poor agreement between retrospective and prospective measures of childhood adversities ^(58)^, retrospective measures have been shown to be valid in population studies, thought they might underestimate the association with objectively measured outcomes, such as CVD ^(60)^. Information on exposure to childhood maltreatment was assessed in a questionnaire completed after information on mediators was collected (mediators were assessed at baseline). However, childhood maltreatment, by definition, related to experiences in early life. The report of childhood maltreatment could plausibly be affected by concurrent mental health, such that individuals who develop depression or report increased psychological distress are more likely to report an adverse childhood experience ^(61)^. Therefore, results found for anxiety/depression could be overestimated, which would, consequently, underestimate the additional contribution of the other mediators. That said, it is unlikely that smoking status, BMI and CRP levels influence the report of childhood maltreatment (independently of mental health), hence reverse causality with these factors is improbable. Nonetheless, comparing and contrasting evidence from studies with both prospective and retrospective measures of childhood maltreatment is recommended.

Measurement error or misclassification of the mediators, particularly smoking, also cannot be ruled out. Ideally, we would have used better data on smoking, such as number of pack-years of smoking, but there was a high proportion of missing data (32%), due to missing on age at which participant started and stopped smoking. Random measurement error or misclassification on the mediators would lead to underestimation of the indirect effect and, therefore, overestimation of the direct effect of childhood maltreatment on CVD^(62)^.

Previous studies have shown that childhood maltreatment is associated with poor adult socioeconomic outcomes ^(16)^, and that adult SEP mediates up to 27% of the association between childhood adversities and health risks, such as depressive disorder, smoking, binge drinking, obesity and sub-optimal health ^(63)^. Therefore, adult SEP should be considered when trying to understand the behavioural, mental health and biological mechanisms through which childhood maltreatment affects CVD, as we were able to do here. Yet bias due to unmeasured confounding cannot be ruled out. We were also able to decompose the total effect into direct and indirect effects even in the presence of exposure-mediator interactions, a limitation of traditional mediation methods ^(10, 64)^. Another strength is that the large sample size enabled us to explore mediating pathways of the associations between different types of childhood maltreatment and CVD, as well as by sex.

The main limitation of this study is the use of a sub-sample of UK Biobank, which itself includes less than 6% of the individuals aged 40-69 years who were invited to enter the cohort, and is not representative of the UK population ^(65)^. This can induce collider bias, which, in this case, would lead to negatively biased associations ^(66)^; therefore, the associations observed here are likely to be underestimated.

Causal mediation analysis relies on several strong assumptions about no confounding between exposure-outcome, exposure-mediator and mediator-outcome associations. We adjusted for several measured confounders in the associations between exposures, mediators and outcome, but bias due to unmeasured confounding in these relationships cannot be ruled out. Although we believe the causal ordering of the mediators used in the analyses is sensible, some associations are likely to be bi-directional. However, results from sensitivity analysis changing the sequential order of the mediators did not substantially differ from the main results.

We did not include diet and physical activity. However, we believe that much of the indirect effect of these factors on the total effect of childhood maltreatment on CVD is likely to be via BMI and/or mental health.

## Conclusions

Anxiety/depression, smoking, BMI and inflammation mediated part of the effect of childhood maltreatment on CVD, varying from 26%-90%, and the contribution of these mediators differed by type of maltreatment and sex. These findings contribute to understanding of how childhood maltreatment might affect CVD and highlight the potential to reduce CVD burden by targeting modifiable mediating factors.

## Data Availability

UK Biobank is an open access resource, and the data reported in this study are available via application to the UK Biobank.

## Ethics approval

UK Biobank received ethical approval from the National Health Service National Research Ethics Service (11/ NW/0382).

## Acknowledgments

This research has been conducted using the UK Biobank Resource (application 19278). We are grateful to all participants who took part in this study.

